# Functional gut microbiome dysbiosis is associated with overall survival after solid organ transplantation

**DOI:** 10.64898/2026.09.04.26362223

**Authors:** Shuyan Zhang, J. Casper Swarte, TransplantLines investigators, Tim J. Knobbe, Kevin Damman, Erik A. M. Verschuuren, Tji C. Gan, Hermie J. M. Harmsen, Vincent E. De Meijer, Hans Blokzijl, Stephan J. L. Bakker, Ranko Gacesa, Rinse K. Weersma, Johannes R. Björk

## Abstract

**Background:** Long-term survival after solid organ transplantation remains limited by infection, graft dysfunction, and systemic complications. Recent evidence suggests that gut dysbiosis, marked by loss of beneficial commensals and expansion of opportunistic pathogens, is associated with increased mortality in solid organ transplant recipients (SOTRs). While specific species have been associated with higher mortality risk or improved survival, the functional mechanisms on the strain level underlying these associations remain unclear. Here, we integrated gene-centric functional profiling with strain-level resolved metagenome-assembled genomes (MAGs) across 1231 gut metagenomes from 1008 SOTRs (580 kidney, 245 liver, 133 lung and 50 heart transplant recipients) and 233 healthy controls from the TransplantLines Biobank and Cohort Study to link gut microbial functions and metabolic modules to their originating MAGs and assess their association with post-transplant overall survival.

**Results:** Functional profiling of the gut microbiome revealed pronounced functional dysbiosis in SOTRs compared with healthy controls, with higher dysbiosis scores significantly associated with increased post-transplant mortality. We identified 97 microbial functions (KEGG Orthologs, KOs) positively associated with impaired survival (hazard ratio >1; FDR < 0.10) and 661 KOs associated with improved survival (hazard ratio <1; FDR < 0.10) after transplantation. KOs associated with excess mortality predominantly encoded functions related to bacterial adhesion, virulence, and stress adaptation. In contrast, protective KOs mapped to 12 highly complete metabolic modules, including shikimate and pyridoxal phosphate biosynthesis. Genome-resolved analysis of 10,397 MAGs revealed that KOs associated with excess mortality were concentrated in members of the Proteobacteria phylum, particularly the *Enterobacteriaceae* family, whereas protective KOs were enriched in members of the Firmicutes A lineage.

**Conclusion:** The functional capacity of the post-transplant gut microbiome differed significantly from that of healthy controls, consistent with a state of post-transplant gut dysbiosis. Notably, functional dysbiosis of the gut microbiome was associated with post-transplant survival. These findings highlight microbial functional potential as a determinant of post-transplant survival and a promising target for microbiome-informed risk prediction and therapeutic intervention.

## Introduction

Solid organ transplantation (SOT) has become a life-saving therapy for patients with end-stage organ failure^1^. However, long-term survival of solid organ transplant recipients (SOTRs) remains a critical clinical concern^2^, largely because of their vulnerability to severe infections, graft dysfunction, and systemic complications^3^. In addition to these clinical challenges, SOTRs often suffer from persistent gut dysbiosis, generally characterized by reduced microbial diversity and expansion of opportunistic pathogens at the expense of beneficial commensals^4-^6. This disturbance is driven by a complex interplay of factors, including chronic immunosuppressive therapy^7^, recurrent antibiotic exposure^8^, and dietary changes^9^.

Growing evidence underscores the clinical relevance of gut dysbiosis in SOTRs, as it has been associated with increased risk of infection^10^, reduced health-related quality of life (HRQoL)^11^ and compromised graft tolerance^12^. Notably, alterations in the gut microbiome have also been linked to increased mortality in both the general population^13^ and allogeneic hematopoietic cell transplant recipients^14^, highlighting the broader relevance of host-microbiome interactions for survival. In SOTRs, long-term survival remains limited, with gut dysbiosis potentially both contributing to and being influenced by post-transplant complications that ultimately affect survival. To explore the relationship between gut microbiome and mortality after SOT, we previously investigated the association between gut dysbiosis and both all-cause and cause-specific mortality in SOTRs from the TransplantLines Biobank and Cohort Study^4,15^. Specifically, we found that several opportunistic pathobionts, such as *Escherichia coli, Enterocloster* (fd. *Clostridium*) *boltae, Clostridium innocuum* and *Hungatella hathewayi* were associated with increased post-transplant mortality risk, whereas higher abundances of gut commensals and short-chain fatty acid-producing bacteria were associated with reduced mortality risk^15^.

These previous studies largely focused on linking taxonomic features to post-transplantation outcomes. However, different strains of the same microbial species can exert different immunomodulatory effects on the host^16–19^. As such, taxonomic profiling alone does not capture strain-level variation or the functional heterogeneity within microbial species, limiting insight into the mechanisms through which the gut microbiome may influence mortality. Shifting the focus toward the functional potential of the gut microbiome, together with the microbial strains and species from which these functions originate, could therefore improve our understanding of gut dysbiosis in general and its associations with clinical outcomes in particular.

In this study, we applied a gene-centric approach to profile and identify gut microbial functions associated with mortality in 1,008 SOTRs and 223 healthy controls from the TransplantLines biobank and cohort study. We then linked these mortality-associated functions to their originating species by mapping gene functions to 10,397 metagenome-assembled genomes (MAGs) reconstructed from the same 1,231 gut metagenomes. By integrating functional and genome-resolved analyses, this study provides mechanistic insight into post-transplantation gut dysbiosis and how the gut microbiome may influence survival after transplantation, and offers a foundation for developing microbiome-targeted strategies to improve long-term outcomes in SOTRs.

## Results

### Characteristics of the solid organ transplant recipients

A total of 1008 SOTRs, recruited in the TransplantLines Biobank and Cohort study, were included in this study and provided a fecal sample at a variable time after transplantation. Of these, 580 were kidney (KTR), 245 liver (LTR), 133 lung (LuTR) and 50 heart transplant recipients (HTR) (Table 1). Across all organ types, the mean (standard deviation [SD]) time since transplantation was 4.0 (6.6) years, 613 recipients (61%) were male, and the mean (SD) age of all recipients was 57 (13.0) years. We also included 223 fecal samples collected before surgery from 223 healthy controls who donated a kidney enrolled in the TransplantLines Biobank and Cohort study as controls in this study.

**Table 1.** Demographics and anthropometrics of this study.

| Parameter | Kidney<br>transplant<br>recipients | Liver<br>transplant<br>recipients | Lung<br>transplant<br>recipients | Heart<br>transplant<br>recipients | Healthy<br>controls |
| --- | --- | --- | --- | --- | --- |
| Number of subjects, *n* | 580 | 245 | 133 | 50 | 223 |
| Age, years (median<br>[IQR]) | 59 [49-66] | 59 [49-66] | 61 [55-65] | 59 [49-68] | 58 [50-66] |
| Male sex, *n* (%) | 364 (62.7) | 150 (61.2) | 69 (51.9) | 30 (60) | 109 (47.5) |
| BMI, kg/m<sup>2</sup> | 27.4 ± 4.7 | 26.8 ± 4.7 | 25.8 ± 4.7 | 25.6 ± 3.7 | 26.4 ± 3.4 |
| Events, *n* | 54 | 17 | 30 | 5 | — |
| Infection-related<br>events, *n* | 16 | 4 | 9 | 0 | — |
| Cardiovascular-related<br>events, *n* | 8 | 2 | 5 | 2 | — |
| Malignancy-related<br>events, *n* | 13 | 4 | 8 | 1 | — |
| Events due to other<br>causes, *n* | 17 | 7 | 8 | 2 | — |

For survival analyses, 863 recipients with ≥12 months of follow-up post-transplantation were included to minimize the influence of early post-transplant mortality and surgery-related complications. During the follow-up period (min=1.4 years and max=31.5 years), a total of 106 participants (54 KTR, 17 LTR, 30 LuTR and 5 HTR) died. Of these, 29 (27%) died due to infection-related mortality, 17 (16%) due to cardiovascular-related mortality, 26 (25%) due to malignancy-related mortality, and 34 (32%) died from other causes.

### Functional dysbiosis is associated with mortality after transplantation

We first analyzed the functional potential of the gut microbiome by focusing on KEGG Orthologs (KOs) which group genes based on functional similarity with each KO representing a conserved molecular function. We characterized variation in KO profiles across samples using Principal Component Analysis (PCA) on CLR-transformed KO relative abundances and found, in line with our previous findings on microbial species^4^, a marked difference between SOTRs and controls (P=0.001, Fig. 1a). To characterize the extent of functional dysbiosis in each recipient’s gut microbiome, we computed a functional dysbiosis score defined as the z-score normalized difference between the KO-based Aitchison distance from each recipient to the average of the functional profile of all controls, and the KO-based Aitchison distance from each recipient to the average functional profile of all SOTRs (Fig. 1a). Next, we analyzed its association with mortality and observed that increased functional dysbiosis was associated with increased mortality after transplantation (Cox proportional hazards model: hazard ratio [HR] 1.25, 95% confidence interval [CI] 1.02 to 1.53, P=0.035; Fig. 1b) adjusting for time since transplantation, age, sex, BMI and medication regimen. The top 10 KOs positively correlated with the functional dysbiosis score were functions associated with stress adaptation (e.g., K11926, K11932), including multidrug efflux (K13639), biofilm formation (e.g., K06080, K11941) and nutrient-acid scavenging (e.g., K09996, K05517), whereas the top 10 negatively correlated KOs included functions related to sporulation (e.g., K07699, K06438), anaerobic energy metabolism (K15022), vitamin and nutrient transport (K16927), and flavonoid metabolism (K00091) (Fig. 1c; See Table S1 for all KOs), functions broadly related to pathways previously identified as characteristic of healthier gut microbiomes^20^.

**Fig. 1.**
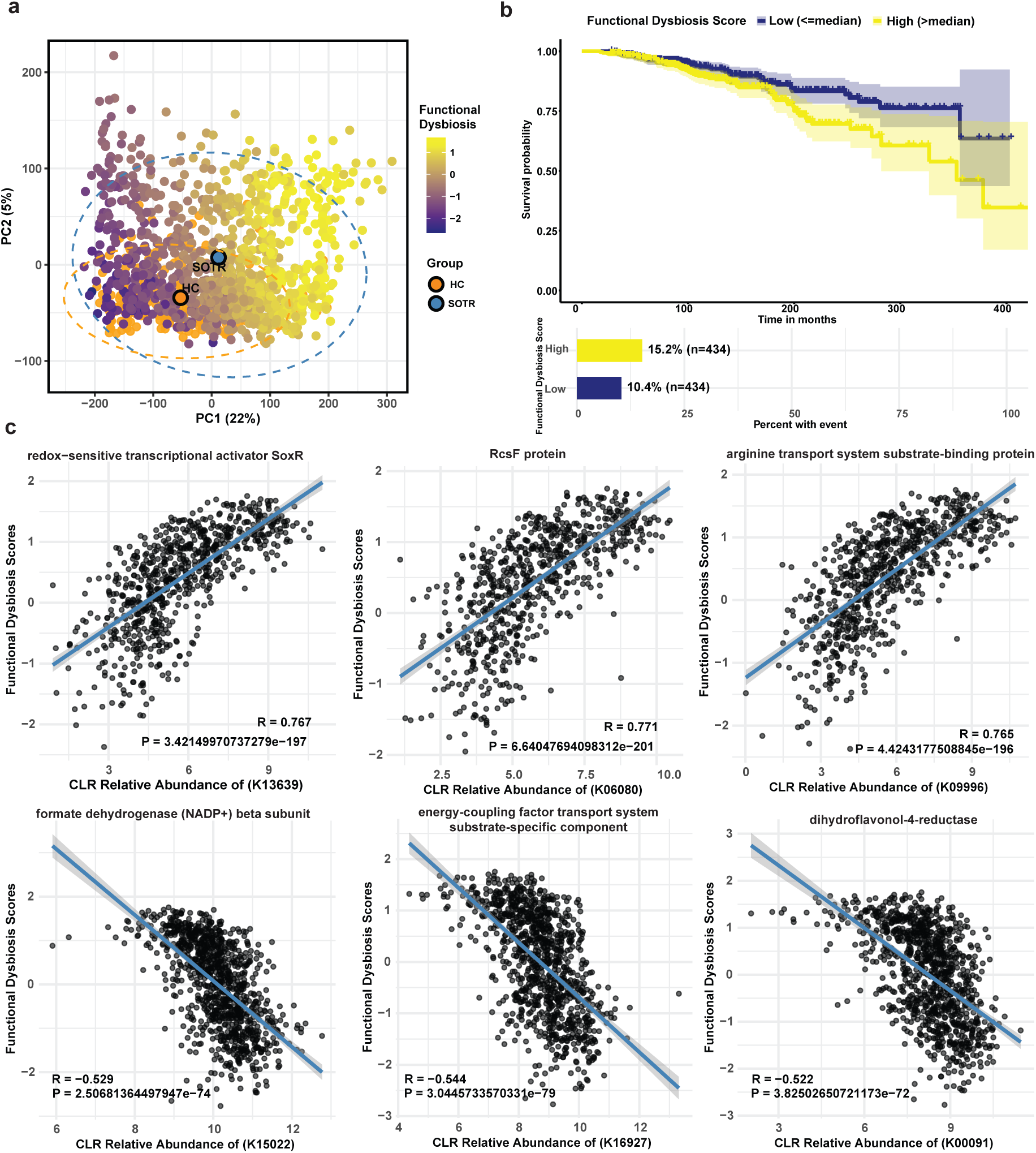
(a) Principal component analysis (PCA) of CLR-transformed KO abundances. Dots correspond to samples with controls in orange and SOTRs colored by the functional dysbiosis score: higher values (more yellow) indicate greater deviation from the controls, whereas lower values (more purple) indicate closer similarity to controls. The large orange and blue circles represent the centroid for the control and SOTR population, respectively. (b) Kaplan-Meier survival curve showing mortality risk in SOTRs stratified by higher (yellow) or lower (purple) than median functional dysbiosis score. High functional dysbiosis score was significantly associated with increased mortality (Cox proportional hazards model, P = 0.035). (c) Pearson correlations between functional dysbiosis scores and CLR-transformed relative abundance of the KOs. Displayed are KOs with the largest absolute effect sizes (|R|).

### Individual functions are associated with all-cause mortality

Using multivariable Cox proportional hazards models, we identified individual KOs associated with mortality within the transplant population. After false-discovery correction (FDR <0.10), we identified 97 (1.55%; Fig. 2a; Table S2) and 661 (10.55%; Table S2) KOs whose CLR-transformed relative abundances were positively and negatively associated with mortality after transplantation, respectively. Reassuringly, 87.6% (85/97) of the KOs associated with increased mortality risk were positively correlated with the dysbiosis score, whereas 99.2% (656/661) of those associated with a decreased mortality risk, were negatively correlated with the dysbiosis score (Table S3).

**Fig. 2.**
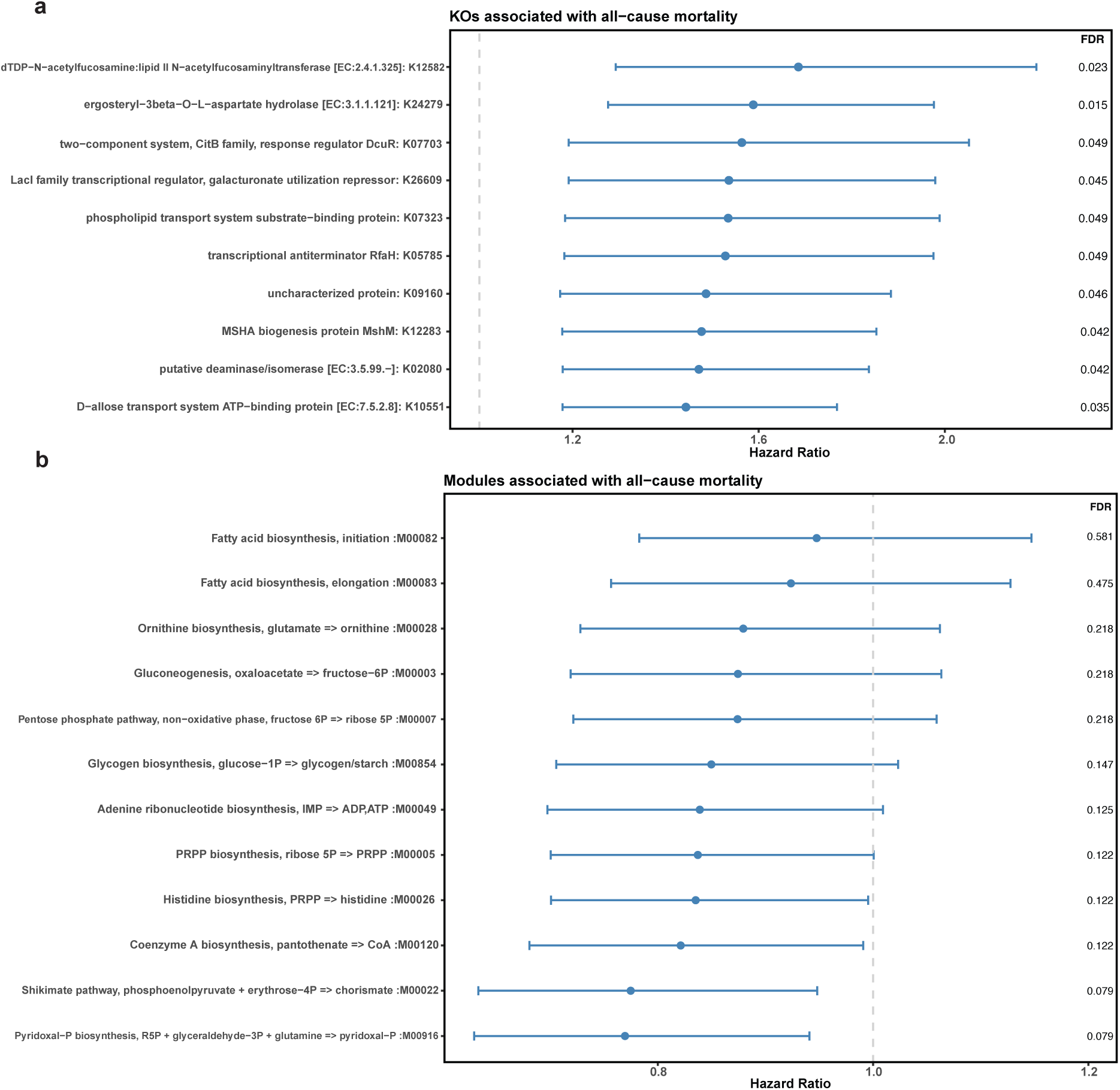
Forest plots of microbial features associated with all-cause mortality in multivariable Cox regression models (a) KOs whose CLR-transformed relative abundances were positively associated with mortality (cutoff: Hazard ratio > 1; FDR < 0.05; for full list, see Table S2). (b) Metabolic modules reconstructed from protective KOs and with high completeness (≥ 75%), whose CLR-transformed relative abundances were associated with lower mortality risk (cutoff: Hazard ratio < 1; FDR < 0.1). Hazard ratios and 95% CIs are shown together with the false discovery rate (FDR)-corrected p values.

Many of the 97 KOs associated with increased mortality risk encoded bacterial functions related to cell-surface structure, including capsule and biofilm components (e.g., K12582, K12283, K12945), peptidoglycan-modifying enzymes (e.g., K14054, K03806, K08308), and flagellar assembly proteins (e.g., K02422, K02394, K02416), consistent with roles in adhesion, persistence, and immune evasion. We also observed multiple virulence-associated genes, including adhesins and fimbrial proteins (e.g., K21964, K19232), siderophore biosynthesis machinery (e.g., K05374), and phage-or competence-associated factors (e.g., K06903, K02241), indicative of enhanced host interaction, iron competition, and horizontal gene transfer. In addition, several KOs reflected stress adaptation, including oxidative-stress enzymes (e.g., K04565, K06879, K17290) and envelope stress regulators and proteases (e.g., K03597, K14061, K01407). Finally, many KOs encoded transporters and efflux systems that enhance nutrient scavenging (e.g., K11737, K10001, K02002), metal and drug resistance (e.g., K07787, K07798, K19273), and metabolic flexibility (e.g., K08484, K10551, K23547). We did not find any associations with cause-specific mortality (Table S4-S7).

We next mapped mortality-associated KOs to KEGG metabolic modules. KEGG metabolic modules are structured sets of KEGG Orthologs (KOs) that together define specific biochemical pathways or functional units, enabling assessment of pathway completeness based on the presence of the constituent KOs. The 97 KOs associated with increased mortality risk mapped to 9 modules, all of which displayed low completeness (pathwise completeness defined as the fraction of steps forming a valid KEGG-defined path supported by the detected KOs) (min=14%, median=16.7%, max=50%). However, the 661 KOs associated with reduced mortality risk mapped to 98 metabolic modules, whereof 12 displayed high completeness (≥ 75%; Fig. S1). Of these, we found that the Shikimate pathway (M00022; 86% completeness) and Pyridoxal phosphate biosynthesis (M00916; 100% completeness) were both significantly associated with decreased mortality risk (FDR<0.1; Fig. 2b).

### Species-level mapping of mortality-associated microbial functions

To identify which species carried the largest number of mortality-associated KOs, we constructed and annotated a total of 10,397 high-quality MAGs (completeness > 90% and contamination < 10%) from the same metagenomes. Across the 254 assigned species, MAGs belonging to the *Enterobacteriaceae* family (phylum Proteobacteria) carried the largest number of mortality-associated KOs (HR>1; Nmin=47, Nmax=70), with *Escherichia coli* MAGs carrying the highest average number (HR>1; Nmean=70; Figure 3, Table S8).

**Fig. 3.**
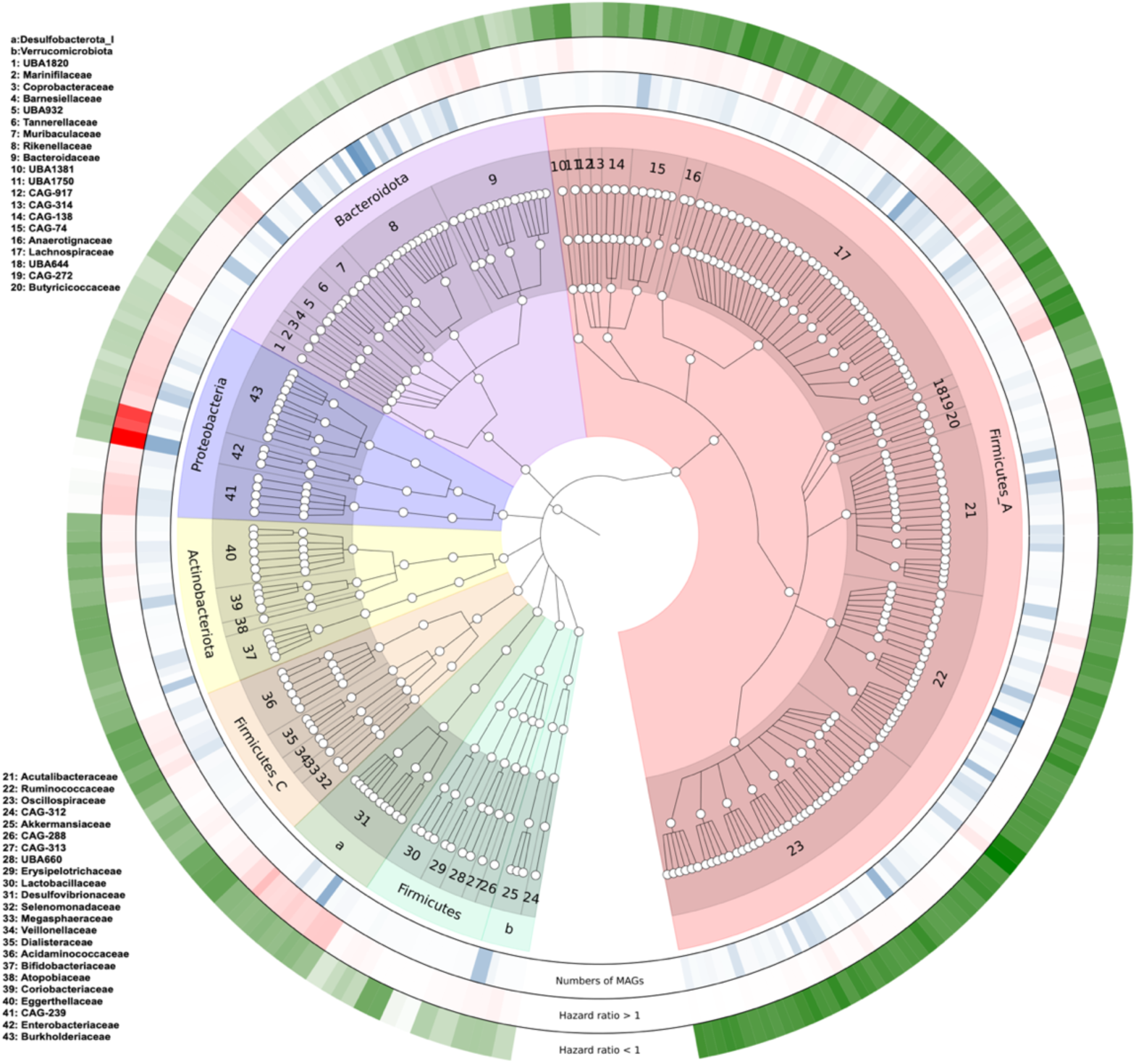
Cladogram depicting high-quality MAGs carrying KOs associated with the all-cause mortality. Each level in the phylogenetic tree corresponds to different taxonomic levels, from domain (the most internal node) to species (the outer tips). Each tip in the phylogeny represents a species for which more than five MAGs were identified, and tips within the same color block represent MAGs from the same microbial phylum. The ring with blue cells shows the number of MAGs mapping to any particular species (range: 7 to 348). The ring with red cells shows the mean number of KOs positively associated with mortality (hazard ratio > 1) carried by MAGs from the same species (range: 1 to 70). Finally, the ring with green cells shows to the average number of KOs associated with reduced mortality risk (hazard ratio < 1) carried by MAGs from the same species (range: 191 to 400). The strength of the ring colors (alpha) was normalized to visualize quantitative differences across species. For the blue ring (number of MAGs per species), was scaled by dividing each value by the maximum. For the red and green rings (mean number of KOs with hazard ratios >1 and <1, respectively), alpha was normalized using min-max scaling so that the minimum and maximum values correspond to the lightest and darkest colors, respectively.

Correspondingly, MAGs from the Proteobacteria phylum carried, on average, the lowest number of KOs associated with decreased mortality risk (HR<1), whereas MAGs from the Firmicutes A (a major Firmicutes lineage in the GTDB taxonomy) carried the highest number of KOs associated with decreased mortality (HR<1; Nmin=272, Nmax=400; Table S8). Notably, half of the top 10 species with the highest average number of KOs associated with survival belonged to the family *Oscillospiraceae*.

### Species abundance does not recapitulate mortality-associated functional associations

We next explored whether higher abundances of MAGs from species harboring a greater number of mortality-associated functions were associated with increased mortality risk after transplantation. To test this, we examined the top 10 species with the highest average number of mortality-associated KOs across their MAGs (Fig. 4). These species belonged to the genera *Desulfovibrio* (Nspecies=1, NMAGs=10), *Enterocloster* (Nspecies=1, NMAGs=12), *Escherichia* (Nspecies=1, NMAGs=222), *Klebsiella* (Nspecies=2, NMAGs=45), *Mailhella* (Nspecies=2, NMAGs=34), *Scatacola_A* (Nspecies=1, NMAGs=63), *Scatocola* (Nspecies=1, NMAGs=57), and *Sutterella* (Nspecies=1, NMAGs=106). A notable feature in Fig. 4 is that genes annotated as tRNA/rRNA methyltransferases (K02533) were present in almost all MAGs from these species except *Enterocloster bolteae*. These enzymes methylate RNA, influencing ribosome function and the efficiency and fidelity of protein translation, particularly under stress conditions such as antibiotic exposure^21^. MAGs from the species *E. coli*, *Klebsiella pneumoniae*, and *K. variicola* not only carried the highest copy numbers of these genes, but also carried the greatest number of distinct mortality-associated KOs overall. However, the relative abundances of MAGs from these species were not significantly associated with mortality after transplantation.

**Fig. 4.**
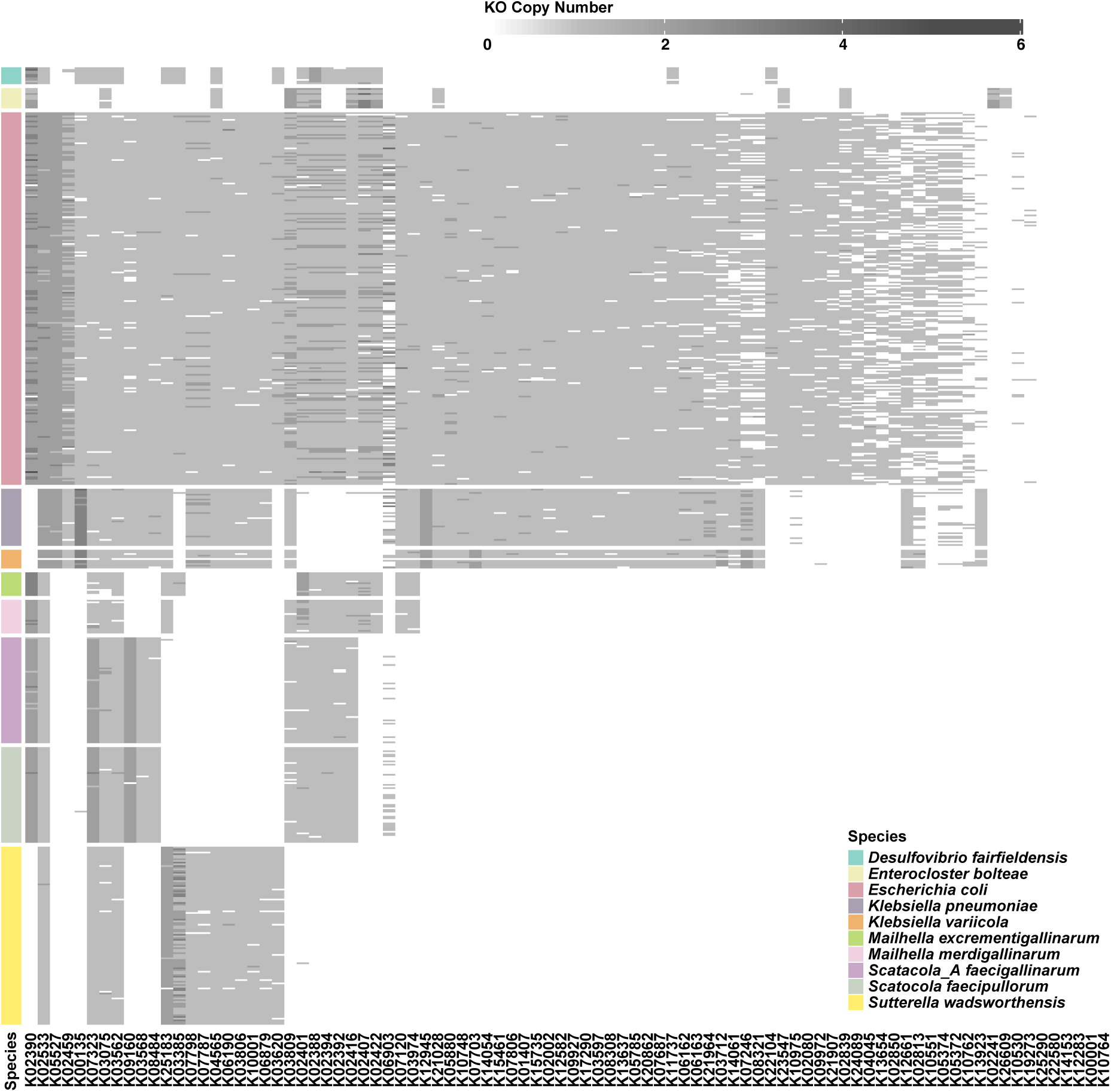
Heatmap showing copy numbers of KOs associated with increased mortality (hazard ratio >1; FDR <0.1) across MAGs for the top 10 species carrying the highest average numbers of such KOs.

Similarly, to link protective microbial functions to specific species, we examined MAGs from species harboring high numbers of KOs associated with decreased mortality risk after transplantation. Species such as *Faecalibacterium prausnitzii* (subspecies C, D, E, and G), *Flavonifractor plautii*, *Gemmiger formicilis*, and *Ruthenibacterium lactatiformans* each harbored, on average, more than 350 of such KOs. Among these species, 353 KOs associated with improved survival were consistently detected in MAGs from all species, including the OmpR-family regulator (K02483), the fatty acid kinase subunit (K25232), and the neurotransmitter sodium symporter (K03308). In contrast, 142 KOs (e.g., aspartate racemase (K01779), energy-coupling factor transport system permease protein (K16925), pyruvate carboxylase (K01958) showed clear strain-specific variation (Fig. 5).

**Fig. 5.**
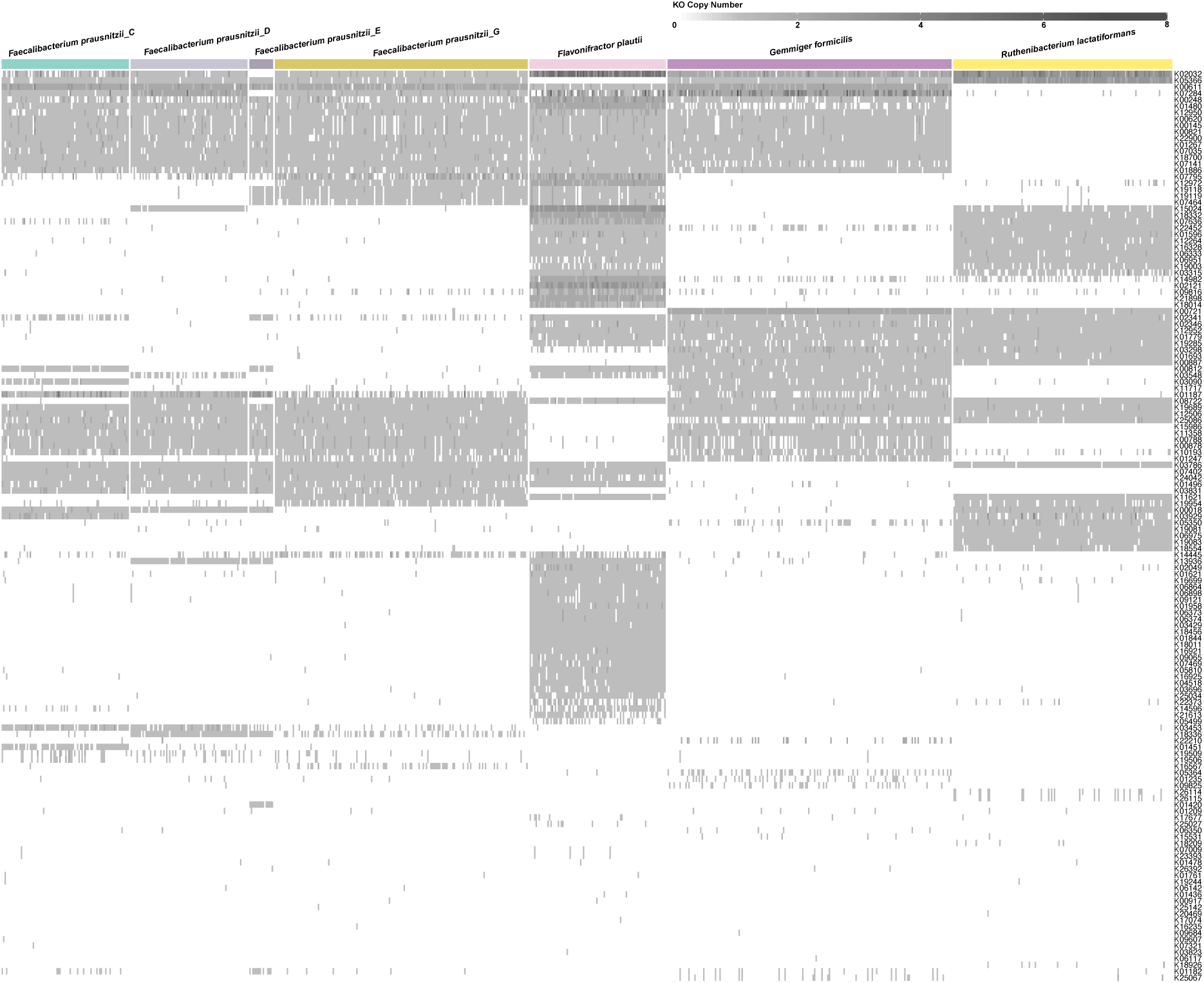
Heatmap showing copy numbers of 142 KOs associated with decreased mortality and exhibiting clear strain-specific variation (hazard ratio <1; FDR <0.1) across MAGs from the species carrying high average numbers (N > 350) of such KOs.

## Discussion

In this study, by integrating functional profiling of microbial genes with metagenome-assembled genomes (MAGs), we linked mortality-associated metabolic functions to their originating strains and species. In total, we identified 97 gut microbial functions associated with increased mortality risk and 661 functions linked to reduced mortality risk. We found that MAGs harboring the highest numbers of mortality-associated functions largely belonged to species previously linked to adverse outcomes after transplantation, whereas those carrying a larger number of functions associated with lower mortality risk were mainly commensals previously associated with host health^11,15^.

Our findings indicate that mortality risk in SOTRs is associated not only with the enrichment of specific bacterial taxa, as previously reported^15^, but also with the functional repertoires some of these taxa harbor. For example, a strong association between members of the *Enterobacteriaceae* family and increased mortality risk was previously demonstrated in a general population over a 15-year follow-up^13^. In our cohort, MAGs identified as *E. coli*, *K. pneumoniae* and *K. variicola*—all members of the *Enterobacteriaceae* family—harbored the highest number of mortality-associated KOs, including the highest copy number of the tRNA/rRNA methyltransferase (K02533). The near-universal presence and high copy number of this KO in mortality-associated species may reflect enhanced translational capacity and stress tolerance, including resilience to antibiotic exposure, traits characteristic of opportunistic, facultative anaerobes that expand under dysbiotic conditions^21,22^.

Moreover, copper/silver efflux systems (e.g., K07787, K07798), which enable bacteria to resist host antimicrobial defenses mediated by copper toxicity^23^, were detected in MAGs from mortality-associated species such as *Sutterella wadsworthensis*, *E. coli*, and other members of the *Enterobacteriaceae* family. Likewise, several transcriptional regulators associated with increased mortality risk, including the TetR/AcrR family regulator (K09017) and the GntR family regulator (K15735), were identified in *Enterobacteriaceae* MAGs. These regulators are known to mediate bacterial adaptation to environmental stress by modulating efflux-mediated antimicrobial resistance, metabolic flexibility, and virulence during inflammation, antibiotic exposure, or nutrient limitation^24,25^.

Together, these mortality-associated functions suggest an enrichment of traits linked to stress adaptation and host interaction, which may contribute to increased mortality risk in SOTRs by promoting opportunistic infections in immunocompromised subjects.

Conversely, we observed a clear enrichment of metabolic functions in gut microbial species associated with lower mortality risk. Half of the 10 species whose MAGs carried the highest numbers of KOs associated with reduced mortality belonged to the *Oscillospiraceae* family, members of which are known for their butyrate-producing and anti-inflammatory properties^26^. MAGs identified as *Gemmiger formicilis* and *Faecalibacterium prausnitzii* also carried high numbers of protective KOs, consistent with their previously reported association with improved post-transplant survival^15^.

Notably, all KOs involved in fatty acid biosynthesis elongation (M00083) and most KOs involved in fatty acid initiation (M00082) were associated with reduced mortality risk, suggesting that microbial lipid biosynthetic capacity may serve as a marker of improved post-transplant outcomes. Lipids play a central role in maintaining bacterial membrane integrity and mediating host-microbe signaling, including immune modulation^27^.

In addition, many protective KOs were involved in the phosphate acetyltransferase-acetate kinase pathway (M00579; acetyl-CoA → acetate). Acetate which is a short-chain fatty acid produced by gut microbes supports gut barrier integrity and modulates immune responses. More broadly, short-chain fatty acids are generally associated with host health and may contribute to reduced mortality risk after transplantation^28^.

Finally, the association between reduced mortality risk and higher abundance of the shikimate pathway (M00022) and pyridoxal phosphate biosynthesis (M00916) suggests that microbial amino acid and vitamin biosynthesis may support immune homeostasis, metabolic stability, and resilience to systemic stress in the post-transplant setting^29,30^.

Interestingly, although MAGs from species such as *E. coli*, *K. pneumoniae*, and *K. variicola* consistently carried large numbers of mortality-associated KOs, the relative abundances of these species were not themselves significantly associated with mortality after transplantation. The largely conserved distribution of mortality-associated KOs across MAGs within individual species suggests that this discrepancy is unlikely to be explained solely by rare strain-specific accessory genes. Instead, these findings indicate that species-level relative abundance may inadequately capture clinically relevant variation in community-wide functional potential. Mortality-associated KOs were distributed across multiple taxa, implying that the cumulative abundance of these functions at the microbiome level may provide a stronger signal than the abundance of any single carrier-species alone. Moreover, the mortality-associated KOs likely constitute only a relatively small fraction of the total genomic content and metagenomic read abundance assigned to these species, potentially weakening associations at the taxonomic level.

Despite the integrative approach combining gene-centric and genome-resolved analyses, several limitations should be noted. Functional annotations based on existing databases may not capture novel or poorly characterized microbial genes relevant to transplant outcomes. In addition, although MAGs provide valuable genome-level resolution, incomplete assemblies may have limited the accurate assignment of certain functions to specific taxa.

Overall, our analyses identify gut microbial functions associated with post-transplant mortality and reveal their distinct distribution across major bacterial lineages through genome-resolved annotation. MAGs belonging to the phylum Proteobacteria harbored the greatest number of mortality-associated KOs and the fewest functions associated with improved overall survival. In contrast, MAGs within the Firmicutes A lineage carried the highest number of protective KOs and relatively few mortality-associated functions. Interestingly, although several species consistently carried large repertoires of mortality-associated KOs, MAG-derived abundances of these species were not themselves significantly associated with mortality. Together, these findings suggest that mortality-associated microbial functions may reflect a broader community-level functional state distributed across multiple taxa, rather than the expansion of individual species alone, and indicate that therapeutic strategies aimed at reshaping microbiome functional composition may represent a possible avenue to improve post-transplant outcomes.

## Conclusion

This study explores functional gut dysbiosis after solid organ transplantation, showing that shifts in the strain-resolved metabolic functional capacity of the gut microbiome are associated with mortality. Mortality-associated functions reflecting enhanced stress tolerance, environmental adaptation, and virulence potential were enriched in Proteobacteria taxa such as *E. coli* and *K. pneumoniae*. In contrast, commensal bacteria within Firmicutes A, including *Faecalibacterium prausnitzii* and members of the *Oscillospiraceae* family, carried high numbers of functions associated with reduced mortality risk, such as KOs associated with gut and immune homeostasis. Together, these findings highlight that the protective or pathogenic potential of the gut microbiome in SOTRs is shaped not only by taxonomic composition but also by its underlying functional repertoire, underscoring the importance of incorporating microbial functional profiling into risk stratification and therapeutic strategies following transplantation.

## Methods

### Study design

We recruited 1008 adult patients who had undergone a kidney (n=580), liver (n=245), lung (n=133), or heart (n=50) transplantation, along with a control group of 223 healthy kidney donors whose fecal samples were collected prior to surgery. As part of the TransplantLines Biobank and Cohort Study (trial registration number NCT03272841, registered on September 6^th^, 2017), the collection of fecal samples, along with extensive phenotype details, have been previously described^31,32^.

### Clinical characteristics

Demographic information and medication use were self-reported and subsequently verified during the study visit. Anthropometric measurements, including height and weight, were obtained by the standardized assessments^31,32^. In this study, the primary clinical outcome was overall survival, using January 1^st^ 2022 as the censoring date. If a patient was deceased, we determined the cause of death and classified it into one of the following categories: cardiovascular, infection, malignancy, or other. At the time of sampling, there were no patients with acute rejection.

### Metagenome-assembled genomes reconstruction

Metagenomic sequencing was performed on fecal DNA samples from 1008 SOTRs and 223 healthy controls. KneadData (v0.5.1)^33^ was used to filter out adapters and low-quality reads (Phred score < 30). Then reads aligned to the human genome (hg19) were removed using Bowtie2 (v2.3.4.1)^34^. Read quality was then assessed by FastQC toolkit (v0.11.7). High-quality clean reads were used for de-novo assembly by MEGAHIT^35^ with a wide range of k-mer sizes (–k-list 21,41,61,81,101,121,141). Assembled contigs (minimum length threshold 1000 bp) were binned using the MetaWRAP binning module with default parameters^36^. These bins were subsequently refined using MetaWRAP’s bin_refinement module to obtain non-redundant bins with ≥ 50% completeness and ≤ 10% contamination. The relative abundance of refined bins was quantified using MetaWRAP’s quant_bins module, which maps cleaned reads back to the refined bins and calculates their relative abundance across samples. The quality of metagenome-assembled genomes (MAGs) was assessed by CheckM’s lineage workflow (lineage_wf)^37^. We retained high-quality MAGs (completeness > 90%; contamination < 10%) for further analyses. Taxonomic classification of MAGs was performed using GTDB-Tk (v2.1.1) with the GTDB-R214 database^38^, and the resulting taxonomy was used to construct a phylogenetic tree using GraPhlAn^39^. Functional annotation and metabolic module reconstruction of the high-quality MAGs were performed using the ‘anvi-estimate-metabolism’ program in anvi’o^40^. This yielded a list of predicted and annotated Kyoto Encyclopedia of Genes and Genomes (KEGG) Orthologs (KOs) for each MAG.

### Functional prediction of metagenomes

Protein-coding sequences (CDSs) were predicted by BAKTA for the assembled contigs from the 1008 SOTRs and 223 healthy controls^41^. CDSs of length ≥300 bp were selected by seqtk to construct a non-redundant gene catalogue using MMseqs2 (parameters: --min-seq-id 0.8-c 0.9)^42^. Using KofamScan^43^, 35.8% of the non-redundant representative genes were assigned to 13,015 unique KOs, with annotations meeting the required e-value ≤ 1e^-10^ and sequence identity ≥ 30%. For genes with multiple KO matches, we retained only the best hit, defined as the match with the lowest e-value and the highest sequence identity. We used bowtie2 to align clean reads of each metagenome to the representative genes, of which the average coverages were then calculated using CoverM^44^. Subsequently, the coverage values of the representative genes annotated to the same KOs were summed to generate KO-level abundance profiles for each metagenome.

We annotated microbial gene functions (KEGG Orthologs, KOs) using the KEGG database (release 111.0), and computed their sample-specific abundances for 1,008 SOTRs and 223 healthy controls, respectively. In total, we identified 13,015 unique KOs from the 1,231 gut metagenomes.

## Statistical analyses

All analyses were performed in R version 4.1, unless stated otherwise.

### Functional Dysbiosis Score

Functional dysbiosis was evaluated based on KOs profiled in 1008 SOTRs and 223 HCs. From the 13,015 detected KOs, identifiers with a prevalence larger than 5% in either SOTRs or HCs were retained, followed by keeping those with a prevalence higher than 20% across the combined dataset, retaining 6,555 KOs for the calculation of the functional dysbiosis score. Centered log-ratio transformation was applied to the relative abundance of the 6,555 KOs and used as an input for the calculation of the Aitchison distance. Functional dysbiosis scores for each SOTR were calculated as the z-score normalized difference between the average Aitchison distance to HCs and the average Aitchison distance to SOTRs.

### Medication regimens

Six different medication regimens of the 1008 transplant recipients were identified by hierarchical cluster analysis using hclust (…,method=”ward.D”) function from the stats package. Medication use was first converted into a binary matrix which was transformed into a dissimilarity object using the vegdist (…,method=”jaccard”) function from the vegan package (Fig. S2).

### Mortality analysis

We applied centered log-ratio (CLR) transformation to relative abundances of all 13,015 KOs in each sample, and then retained 6,265 KOs with a prevalence greater than 50% across SOTR samples collected at least 12 months after transplantation, a time point chosen to reduce surgery-related mortality. To analyze the association between CLR transformed KO abundances and mortality, we used multivariable Cox proportional hazard models as implemented in the function *coxph()* in the R package survival (V.3.5-5), adjusting for age, sex, BMI, months since transplantation and medication regimens, with false discovery rate (FDR) correction of 0.1. The proportional hazards assumption for each included covariate was evaluated using Schoenfeld residuals (P > 0.05). We performed Kaplan-Meier survival analyses by stratifying recipients based on whether the relative abundance of the focal feature was above or below the median.

## Ethics approval

All participants signed an informed consent form prior to sample collection. TransplantLines (METc 2014/077) and Lifelines (METc 2017/152) were approved by the local institutional ethics review board (IRB) from the UMCG. Both studies adhere to the UMCG Biobank Regulation and are in accordance with the World Medical Association (WMA) Declaration of Helsinki and the Declaration of Istanbul.

## Data availability

Data are available on reasonable request. The raw microbiome sequencing data and basic phenotypes used in this study are available at the European Genome-Phenome Archive under accession numbers EGAD00001008907 (https://ega-archive.org/datasets/EGAD00001008907), EGAS00001006257 (https://ega-archive.org/studies/EGAS00001006257) and EGAS00001006258 (https://ega-archive.org/studies/EGAS00001006258). Due to patient confidentiality, the clinical datasets associated with the metagenomic datasets are available on request to the University Medical Centre Groningen. Access to this clinical dataset requires a minimal access procedure consisting of a request per email for a data access form. A response will be provided within two working weeks. This access procedure is to ensure that the data are being requested for research/scientific purposes only and thus comply with the informed consent signed by TransplantLines participants, which specifies that the collected data will not be used for commercial purposes.

## Code availability

All analyses were performed using publicly available software and packages as described in the Methods. The scripts used for the bioinformatics processing and statistical analyses will be made available to the editors and referees upon request.

## Acknowledgements

We would like to thank the Centre for Information Technology of the University of Groningen (RUG) for their support and for providing access to the Hábrók high-performance computing cluster. The TransplantLines Biobank and Cohort study received funding from Astellas BV (TransplantLines Biobank and Cohort study) and Chiesi Pharmaceuticals BV (PA-SP/PRJ-2020-9136) and was co-financed by the Dutch Ministry of Economic Affairs and Climate Policy by means of the PPP-allowance made available by the Top Sector Life Sciences & Health to stimulate public-private partnerships.

R.K.W. and J.R.B are supported by the Seerave Foundation, and R.K.W is also supported by the Netherlands Organization for Scientific Research (NWO), and the EU Horizon Europe Program grant miGut-Health: personalized blueprint of intestinal health (101095470). The funders played no role in the study design, data collection, analysis, reporting, or the decision to submit for publication.

## Author contributions

S.Z. preformed all the analyses with supervision from J.R.B.. S.Z. and J.R.B. drafted the first version of the manuscript, and S.Z., J.R.B., and R.K.W. finalized subsequent versions of the manuscript. All authors critically revised and approved the final version of the manuscript. J.C.S., TransplantLines investigators, T.J.K., K.D., E.A.M.V., C.T.G, H.J.M.H, V.E.de M, H.B., S.J.L.B and R.G. collected data and assisted in study planning.

### Members of TransplantLines Investigators

Coby Annema, Stephan JL Bakker, Stefan P Berger, Hans Blokzijl, Frank AJA Bodewes, Marieke T de Boer, Kevin Damman, Martin H de Borst, Arjan Diepstra, Gerard Dijkstra, Rianne M Douwes, Caecilia SE Doorenbos, Michele F Eisenga, Michiel E Erasmus, C Tji Gan, Antonio W Gomes Neto, Eelko Hak, Bouke G Hepkema, Frank Klont, Tim J Knobbe, Daan Kremer, Henri GD Leuvenink, Willem S Lexmond, Vincent E de Meijer, Hubert GM Niesters, Gertrude J Nieuwenhuis-Moeke, L Joost van Pelt, Robert A Pol, Robert J Porte, Adelta V Ranchor, Jan Stephan F Sanders, Marion J Siebelink, Riemer JHJA Slart, J Casper Swarte, Daan J Touw, Marius C van den Heuvel, Coretta van Leer-Buter, Marco van Londen, Erik AM Verschuuren, Michel J Vos, Rinse K Weersma.

## Supplementary figures

**Fig. S1.**
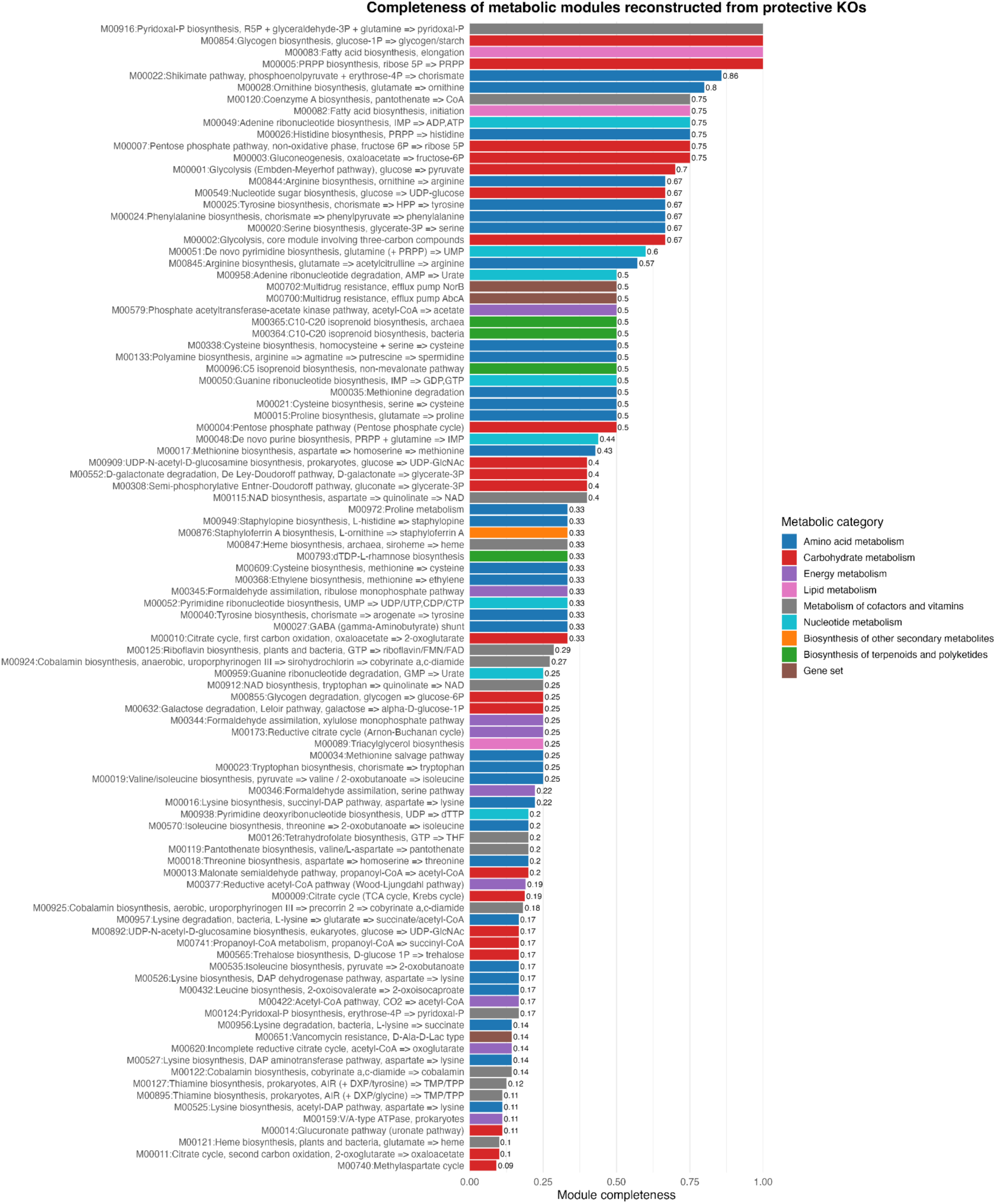
Completeness of metabolic modules reconstructed from KOs associated with lower mortality risk.

**Fig. S2.**
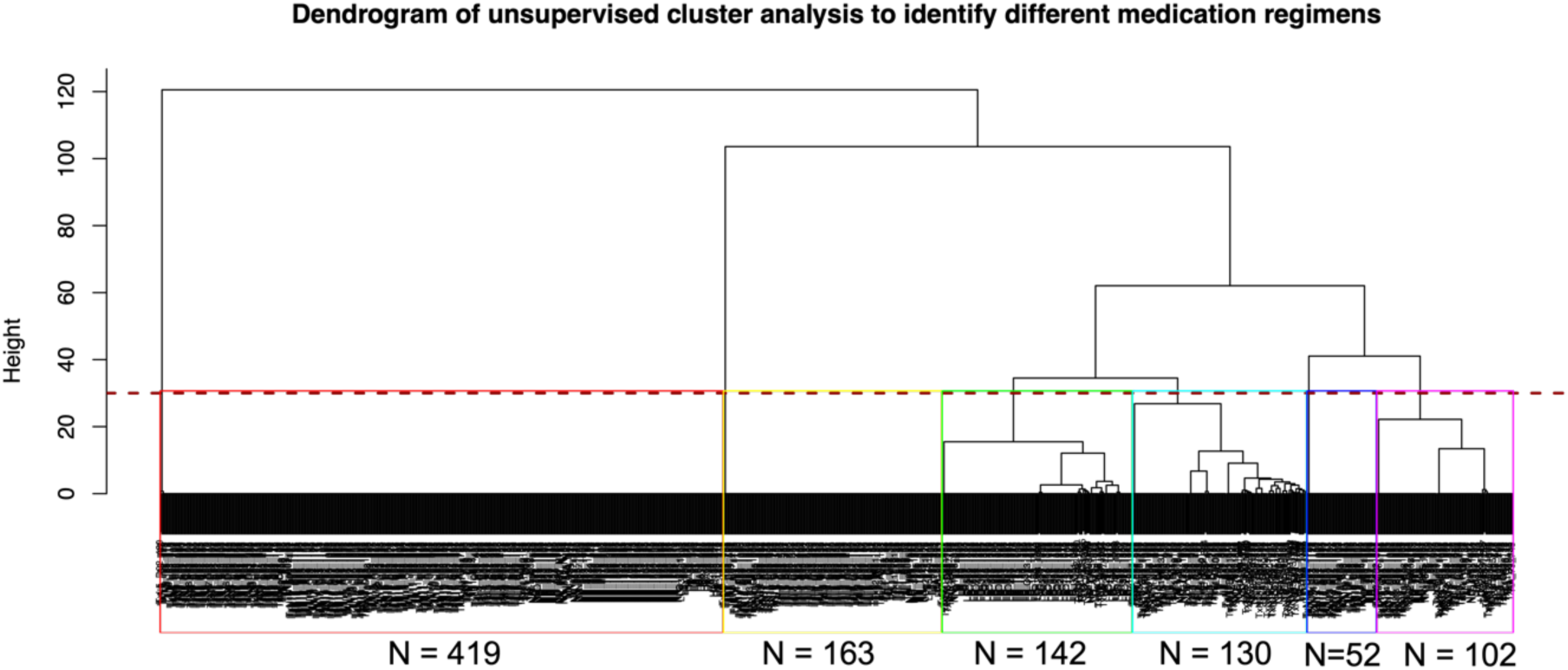
Dendrogram of unsupervised cluster analysis to identify different medication regimens. Dendrogram shows the hierarchical relationship among samples from solid organ transplant recipients. The height at which two samples are joined together represents the dissimilarity of their medication regimens. The larger the height, the more different the medication regimens. Samples joined together at a height of zero were on the same medication regimen. The dendrogram was cut at the height of 30, and each medication regimen includes more than 50 recipients (different colour rectangles) for further analyses.

